# Impact of Hypertension on Left Heart Remodelling: Echocardiography and Computed Tomography Study

**DOI:** 10.1101/2023.06.27.23291981

**Authors:** Aleksandra Lange, Viktoria Palka, Chris Bian, Harry Huntress, Jill Morgan, Sean Allwood, Rohan Swann, Przemysław Palka

## Abstract

**Background:** Early recognition of left atrial (LA) and left ventricular (LV) remodelling may improve prognosis of patients with hypertension.

**Methods:** Left heart coupling indices were studied to investigate the adaptive effect of hypertension on cardiac remodelling. The ratio of LV to LA volumes was measured at selected phases of a cardiac cycle using computed tomography (CT) coronary angiography and transthoracic echocardiogram (TTE). A group of 180 patients was divided into Group 1 (no hypertension) and Group 2 (hypertension). Volume ratios were measured in diastasis by both CT and TTE: LV_dias_ and LA_dias_. Using TTE, volumes were measured at end-diastole (LV_ED_ and LA_min_)), and end-systole (LV_ES_ and LA_max_). LA function and LV/LA ratios were analysed at: LV_ED_/LA_max_, LV_dias_/LA_dias_, LV_ED_/LA_min_, LV_ES_/LA_min_.

**Results:** There were no differences between the age groups, LV_ED_, and LV mass index. Compared to Group 1, Group 2 had bigger mean LV wall thickness (0.90±0.16 cm *vs* 0.83±0.14 cm; p=0.006), increased relative wall thickness ratio (0.39±0.09 *vs* 0.35±0.008, p=0.004), and more frequent concentric LV remodelling (31 *vs* 12, p=0.020). LA volumes were bigger in Group 2 for LA_max_, LA_dias_, and LA_min_. All LV/LA volume ratios were lower in Group 2 compared to Group 1 (LV_ED_/LA_max_ 1.55±0.56 *vs* 1.79±0.69, p=0.010; LV_ED_/LA_min_ 3.56±2.00 *vs* 4.59±2.56, p=0.003; LV_ES_/LA_min_ 1.36±0.77, p=0.005, LV_dias_/LA_dias_ (TTE) 1.75±0.61 *vs* 2.24±1.24, p<0.001, LV_dias_/LA_dias_ (CT) 1.49±0.23 *vs* 1.69±0.41, p<0.001). LA reservoir function and conduit function indexed by E/e’ ratio was lower in Group 2. Combined TTE parameters of relative wall thickness >40, LV_dias_/LA_dias_≤1.81, and indexed by E/e’ LA reservoir function ≤0.068 had the highest discriminate power to differential patients from Group 1 and Group 2 (area under the curve 0.737).

**Conclusions:** In hypertension, prior to the development of LV hypertrophy, adaptive remodelling is based on reduced LV/LA volume ratio, reduced indexed reservoir LA function, and increased relative LV wall thickness.

## INTRODUCTION

Between 1990 and 2019, the number of people with hypertension doubled from 648 million to 1.277 billion worldwide.^1^ Although treatment and control rates have improved, more timely detection of hypertension is needed as the cumulative effect of hypertension ultimately leads to cardiac overload, remodelling, and heart failure (HF).^2–4^ Each month of active antihypertensive therapy is associated with 1-day prolongation of life expectancy free from cardiovascular death.^5^ While it is clear that hypertension increases all-cause mortality, the detection of early adaptive stages of cardiac remodelling is not straightforward.^3, 6–10^ Current criteria focuses on the effects of the maladaptive process, assessed at the point when the left ventricular (LV) wall thickness and mass^11, 12^ are already increased, and LV filling has changed with additional dilatation of both, the left atrium (LA) and the LV.^6, 9^ It has been shown that the best prognosis for patients is during the adaptive stage.^13, 14^ By proposing an easily obtainable score for the early detection of cardiac remodelling, during the adaptive response and prior to the development of LV hypertrophy and LA dysfunction, we would facilitate early diagnosis and better prognosis. We sought to test the clinical significance of the LV to LA volume relationship to address the concept of atrioventricular interplay and changes in hypertensive heart disease in patients with no underlying ischaemic heart disease.^15, 16^ The assessment of LV to LA volume ratio was measured at selected phases of the cardiac cycle using transthoracic echocardiography (TTE) and computed tomography (CT) coronary angiography and modelled as a surrogate for global (net) left-heart atrio-ventricular adaptive remodelling in patients with hypertension.

## METHODS

Our study population was formed of patients aging between 40 to 80 years who were referred for CT coronary angiography between February 2019 to August 2021. Patients with recent onset of shortness of breath and/or chest pain and a low-moderate risk were chosen from our clinical referral cohort if they met the following selection criteria: (1) were in sinus rhythm, (2) had absence of valvopathy ≥2/4, (3) LV ejection fraction was ≥50%, (4) had CT coronary angiography and TTE of diagnostic image quality. Patients with documented myocardial ischaemia and/or history of previous myocardial infarction, coronary artery revascularisation were not included. An original group of 232 patients was selected. From this group, a total of fifty-one patients were excluded. Nine of the fifty-one patients were excluded in view of ongoing symptoms and subsequently documented myocardial ischemia requiring coronary artery revascularisation. Thirty of the fifty-one patients were excluded as their CT coronary angiography was a systolic scan. Twelve of the fifty-one patients were excluded in view of incomplete TTE or CT data (**Figure 1**). This final study group of one-hundred-eighty patients was divided into two groups according to clinical information. Group 1 contained patients who had no clinical evidence of hypertension and diabetes (n=83), and Group 2 had patients diagnosed with hypertension (n=97). There were no differences between Group 1 and Group 2 in relation to basic characteristics including age, gender, and heart rate, **Table 1**. A group of 136 out of 180 (76%) patients were willing to participate in a follow up survey post CT coronary angiography. The average time of the survey was 422±250 days [range 106-1148 days]. Ongoing chest pain was more frequently noted in Group 2 compared to Group 1. No difference was noted between the groups in all other symptoms that included the presence of fatigue, shortness of breath and depression/anxiety.^17^ The quality-of-life score (EQ-5D-5L)^18^ was similar in both groups. Only two major cardiovascular events were noted at the time of the follow up. In Group 1 – one patient had pulmonary embolism diagnosed at day 451, and in Group 2 – one patient required percutaneous coronary artery intervention at day 814.

**Figure 1.**
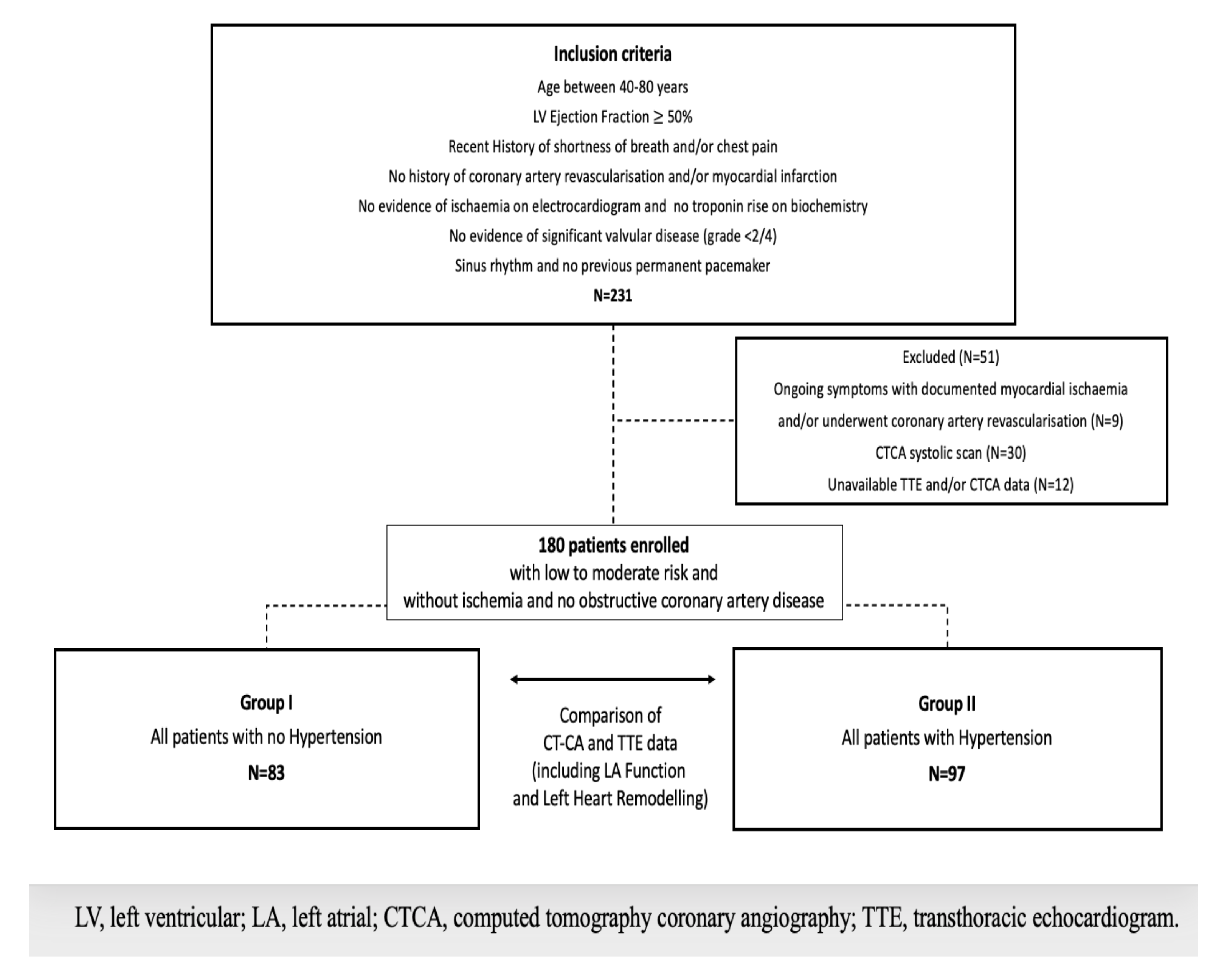
Patients’ selection.

**Table 1.** Patient clinical characteristics and follow-up survey.

| Mean ±Standard deviation or number (%) | Group 1<br>(No hypertension)<br>n=83 | Group 2<br>(Hypertension)<br>n=97 | Correlation<br>coefficient | 95%<br>Confidence interval | P value |
| --- | --- | --- | --- | --- | --- |
| Age (years) | 59±10 | 61±8 | 0.115 | -0.032-0.257 | 0.126 |
| Female gender | 42(51) | 55(56) | 0.061 | -0.086-0.205 | 0.416 |
| Blood pressure, systolic (mmHg) | 125±12 | 138±15 | 0.418 | 0.289-0.531 | <0.001 |
| Blood pressure, diastolic (mmHg) | 75±9 | 80±9 | 0.269 | 0.128-0.400 | <0.001 |
| Heart rate (beats/min) | 66±13 | 67±11 | 0.057 | -0.090-0.201 | 0.449 |
| Obstructive sleep apnoea | 25(30) | 38(40) | 0.095 | -0.052-0.238 | 0.206 |
| Hyperlipidaemia | 44(53) | 67(69) | 0.167 | 0.019-0.304 | 0.027 |
| Type II diabetes | - | 9(9) | 0.212 | 0.068-0.348 | 0.004 |
| Follow-up participants | 58(70) | 78(80) | 0.122 | -0.025-0.264 | 0.102 |
| Days since enrolment<br>[range] | 463±263<br>106-1148 | 392±237<br>92-1114 | -0.141 | -0.302-0.028 | 0.101 |
| Chest pain | 1(2) | 10(13) | 0.223 | 0.038-0.393 | 0.019 |
| Shortness of breath | 8(14) | 15(19) | 0.083 | -0.106-0.266 | 0.391 |
| Depression anxiety stress scale score-21* | 9±8 | 9±7 | -0.005 | -0.192-0.182 | 0.956 |
| Quality of life score† | 75±20 | 78±18 | 0.065 | -0.124-0.249 | 0.503 |

\*, DASS-21<sup>17</sup>; †, EuroQol 5 dimensions 5 levels<sup>18</sup>

The data was prospectively collected and analysed. The study complies with the Declaration of Helsinki. The study was approved by UnitingCare Health Human Research Ethics Committee (number 2019.29.307), Brisbane, Australia. Informed consent has been obtained from all patients. We followed the STARD guidance for conducting and reporting quality.

### TTE

was acquired using commercially available equipment Phillips EPIQ CVx (Philips North America Corporation, Andover, MA) or Siemens SC200 (Siemens Medical Solutions, Mountain View, Calif). Standard clinical imaging protocol was applied to each patient. The protocol consisted of M-mode, 2-Dimensional and Doppler analysis.^19, 20^ Apical views were optimised to avoid foreshortening of the either LV and/or LA. The LV volume was measured using the biplane Simpson’s method at end-diastole – LV_ED_, end-systole – LV_ES_, and at diastasis – LV_dias_. The LA area was planimetered from the 4-chamber and the 2-chamber view, excluding the LA appendage and the pulmonary veins. Subsequent LA volumes were calculated using the biplane area-length method.^19^ LA volume was measured in three phases of the cardiac cycle: maximum – LA_max_ (at LV end-systole), minimum – LA_min_ (at LV end-diastole) and during diastasis – LA_dias_ as previously described.^21^ Mitral inflow peaks early (E) and late diastolic (A) velocities were measured by pulsed wave Doppler. Septal and lateral LV annular velocities (e’) as well as right ventricle lateral annular velocities (s’) were measured using tissue Doppler. Each measurement was averaged from three cardiac cycles. All study patients were divided according to American Society of Echocardiography/European Association of Cardiovascular Imaging recommendations for the classification of LV diastolic dysfunction.^20^ In the indeterminate subgroup, if the difference in pulmonary venous flow atrial reversal duration to mitral inflow A wave duration was more than 30 ms, the data was used as a marker of diastolic dysfunction and elevated LV filling pressure. **Table 2** lists all measured TTE parameters that were taken for the analysis. LA function parameters include: LA reservoir function: total emptying volume and emptying fraction; LA conduit Function: LA passive emptying volume and emptying fraction; LA pump function: LA active emptying volume and emptying fraction.^21^

**Table 2.**
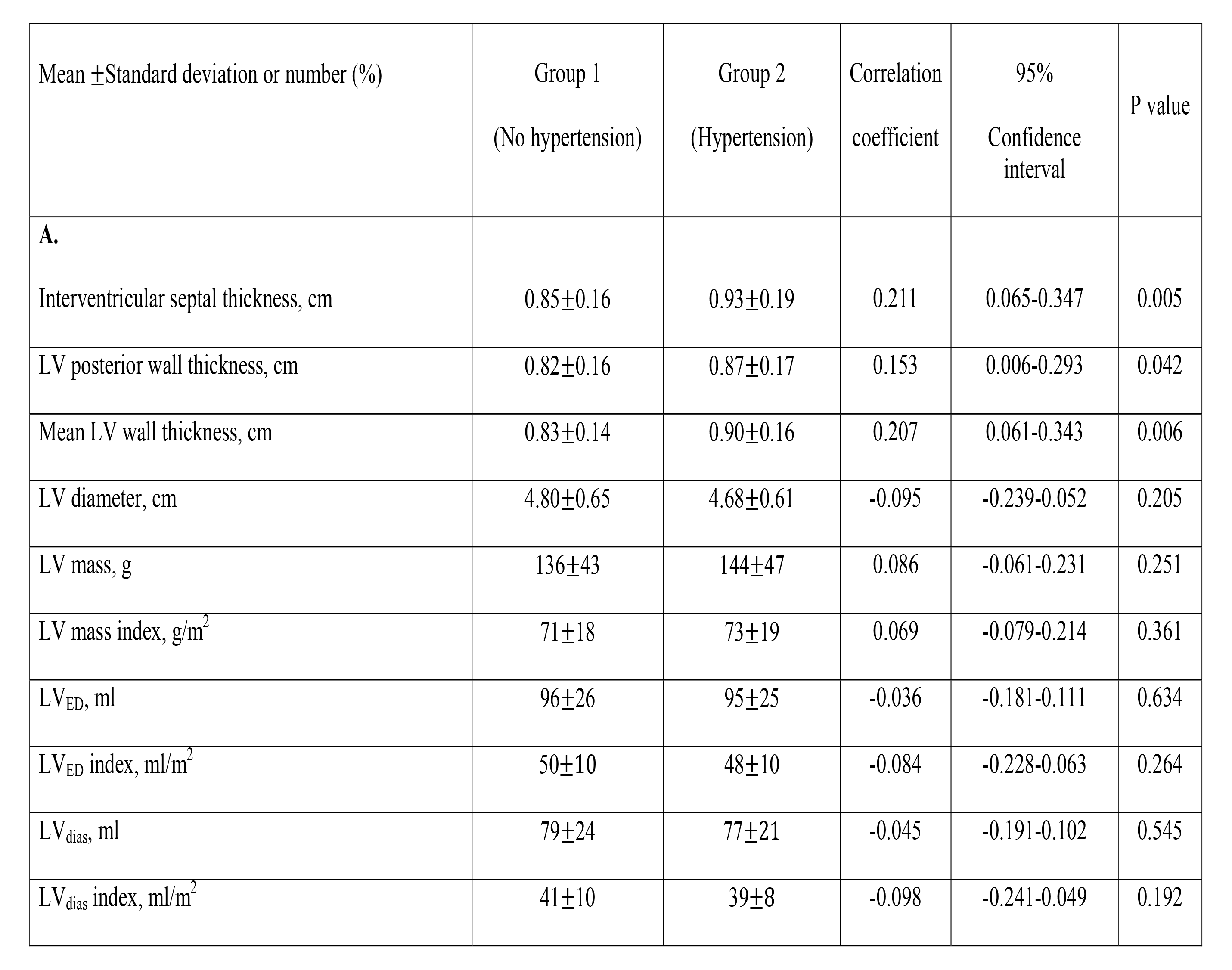
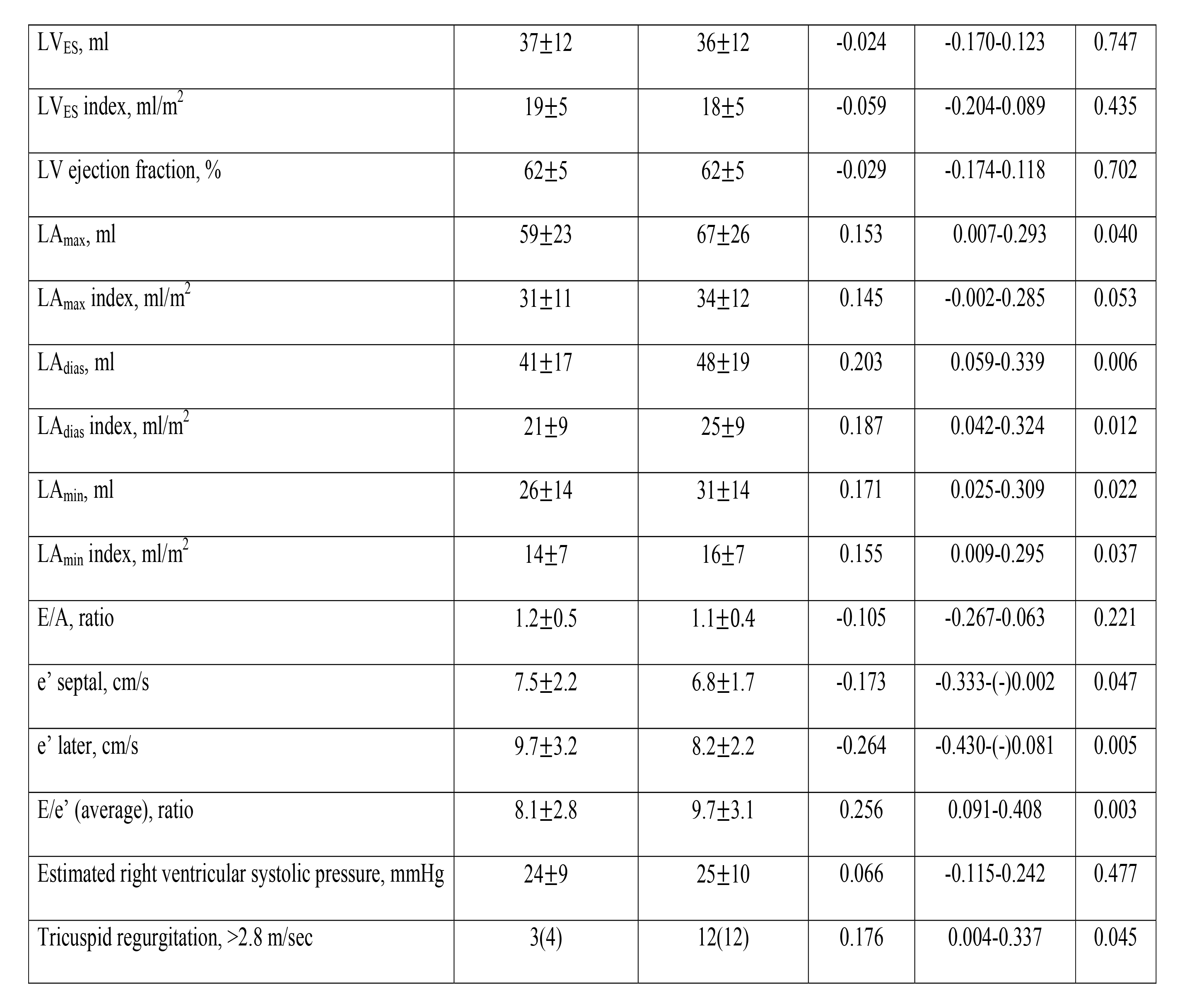
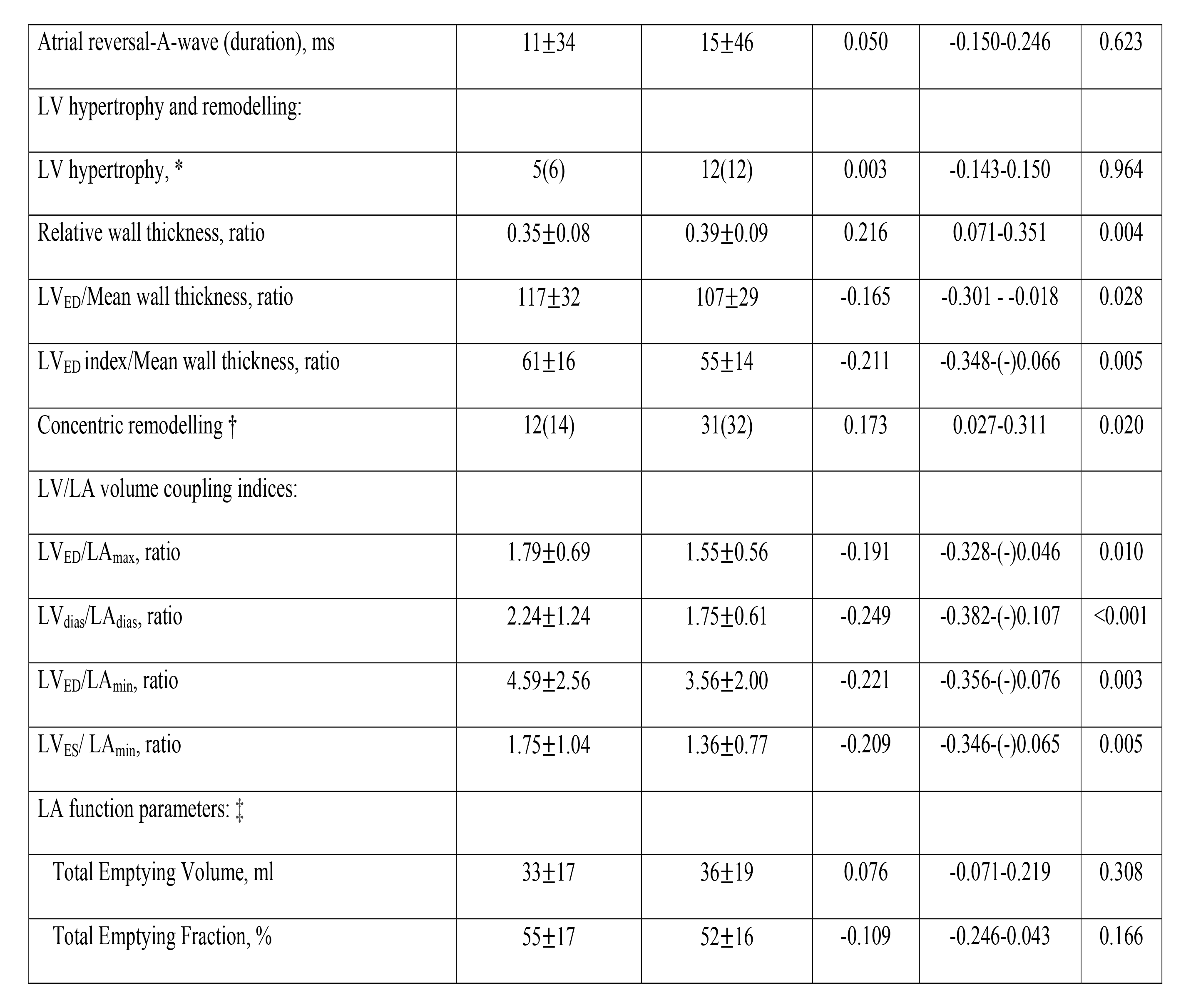
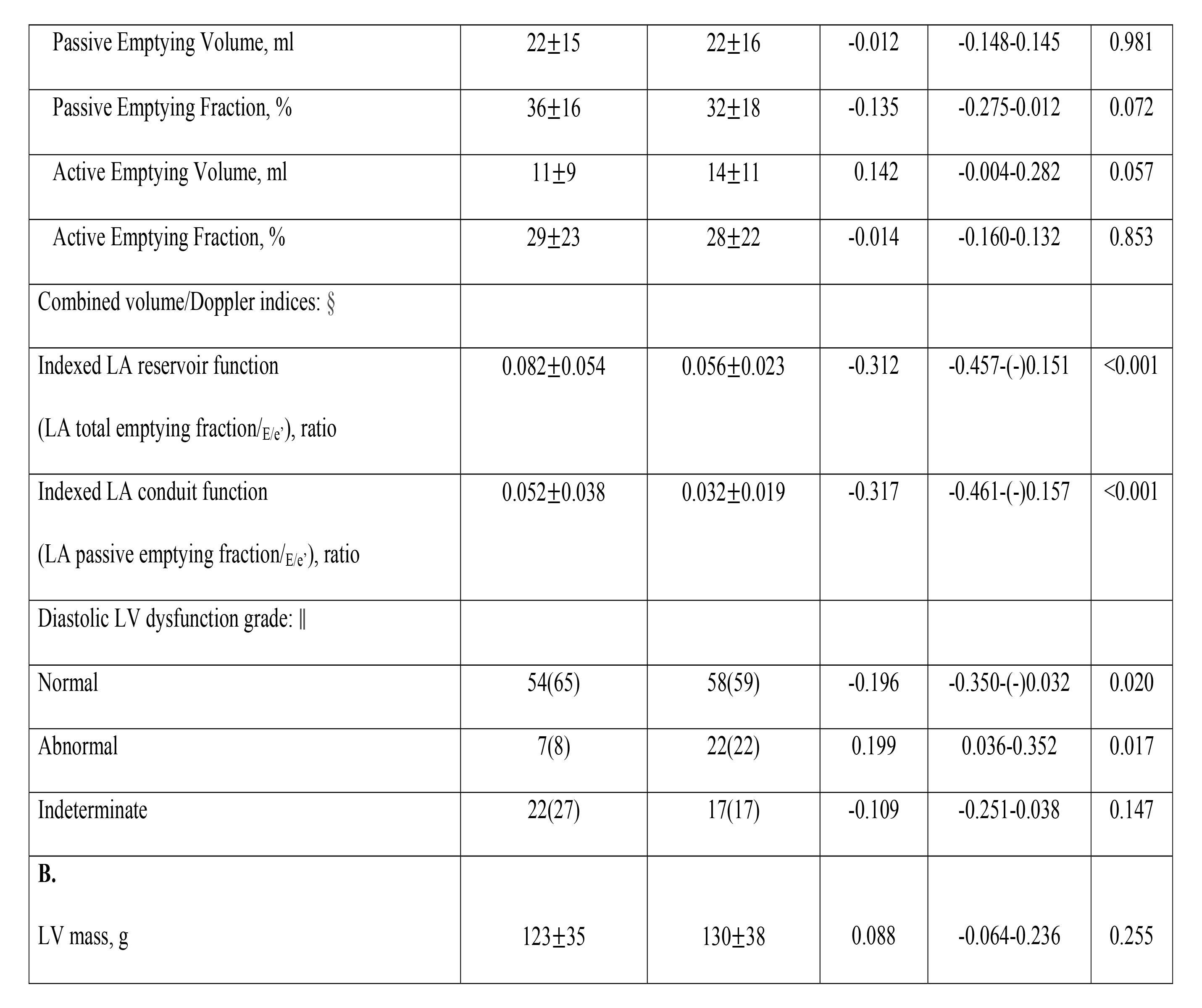
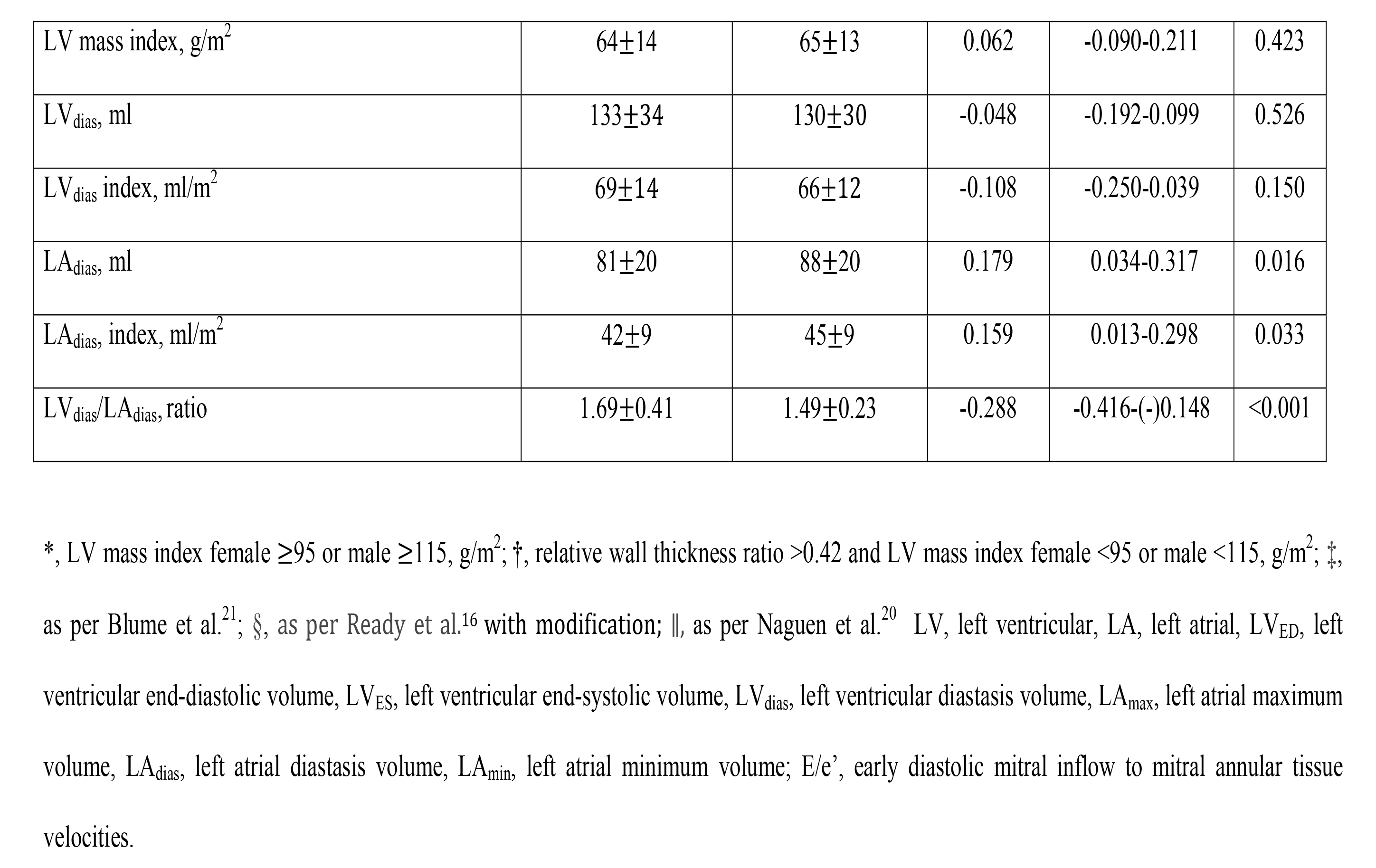
Echocardiographic (**A**) and computed tomography (**B**) data.

| Mean ±Standard deviation or number (%) | Group 1<br>(No hypertension) | Group 2<br>(Hypertension) | Correlation<br>coefficient | 95%<br>Confidence<br>interval | P value |
| --- | --- | --- | --- | --- | --- |
| **A.** |  |  |  |  |  |
| Interventricular septal thickness, cm | 0.85±0.16 | 0.93±0.19 | 0.211 | 0.065-0.347 | 0.005 |
| LV posterior wall thickness, cm | 0.82±0.16 | 0.87±0.17 | 0.153 | 0.006-0.293 | 0.042 |
| Mean LV wall thickness, cm | 0.83±0.14 | 0.90±0.16 | 0.207 | 0.061-0.343 | 0.006 |
| LV diameter, cm | 4.80±0.65 | 4.68±0.61 | -0.095 | -0.239-0.052 | 0.205 |
| LV mass, g | 136±43 | 144±47 | 0.086 | -0.061-0.231 | 0.251 |
| LV mass index, g/m<sup>2</sup> | 71±18 | 73±19 | 0.069 | -0.079-0.214 | 0.361 |
| LV<sub>ED</sub>, ml | 96±26 | 95±25 | -0.036 | -0.181-0.111 | 0.634 |
| LV<sub>ED</sub> index, ml/m<sup>2</sup> | 50±10 | 48±10 | -0.084 | -0.228-0.063 | 0.264 |
| LV<sub>dias</sub>, ml | 79±24 | 77±21 | -0.045 | -0.191-0.102 | 0.545 |
| LV<sub>dias</sub> index, ml/m<sup>2</sup> | 41±10 | 39±8 | -0.098 | -0.241-0.049 | 0.192 |
| LV<sub>ES</sub>, ml | 37±12 | 36±12 | -0.024 | -0.170-0.123 | 0.747 |
| LV<sub>ES</sub> index, ml/m<sup>2</sup> | 19±5 | 18±5 | -0.059 | -0.204-0.089 | 0.435 |
| LV ejection fraction, % | 62±5 | 62±5 | -0.029 | -0.174-0.118 | 0.702 |
| LA<sub>max</sub>, ml | 59±23 | 67±26 | 0.153 | 0.007-0.293 | 0.040 |
| LA<sub>max</sub> index, ml/m<sup>2</sup> | 31±11 | 34±12 | 0.145 | -0.002-0.285 | 0.053 |
| LA<sub>dias</sub>, ml | 41±17 | 48±19 | 0.203 | 0.059-0.339 | 0.006 |
| LA<sub>dias</sub> index, ml/m<sup>2</sup> | 21±9 | 25±9 | 0.187 | 0.042-0.324 | 0.012 |
| LA<sub>min</sub>, ml | 26±14 | 31±14 | 0.171 | 0.025-0.309 | 0.022 |
| LA<sub>min</sub> index, ml/m<sup>2</sup> | 14±7 | 16±7 | 0.155 | 0.009-0.295 | 0.037 |
| E/A, ratio | 1.2±0.5 | 1.1±0.4 | -0.105 | -0.267-0.063 | 0.221 |
| e' septal, cm/s | 7.5±2.2 | 6.8±1.7 | -0.173 | -0.333-(-)0.002 | 0.047 |
| e' later, cm/s | 9.7±3.2 | 8.2±2.2 | -0.264 | -0.430-(-)0.081 | 0.005 |
| E/e' (average), ratio | 8.1±2.8 | 9.7±3.1 | 0.256 | 0.091-0.408 | 0.003 |
| Estimated right ventricular systolic pressure, mmHg | 24±9 | 25±10 | 0.066 | -0.115-0.242 | 0.477 |
| Tricuspid regurgitation, >2.8 m/sec | 3(4) | 12(12) | 0.176 | 0.004-0.337 | 0.045 |
| Atrial reversal-A-wave (duration), ms | 11±34 | 15±46 | 0.050 | -0.150-0.246 | 0.623 |
| LV hypertrophy and remodelling: |  |  |  |  |  |
| LV hypertrophy, * | 5(6) | 12(12) | 0.003 | -0.143-0.150 | 0.964 |
| Relative wall thickness, ratio | 0.35±0.08 | 0.39±0.09 | 0.216 | 0.071-0.351 | 0.004 |
| LV<sub>ED</sub>/Mean wall thickness, ratio | 117±32 | 107±29 | -0.165 | -0.301 - -0.018 | 0.028 |
| LV<sub>ED</sub> index/Mean wall thickness, ratio | 61±16 | 55±14 | -0.211 | -0.348-(-)0.066 | 0.005 |
| Concentric remodelling † | 12(14) | 31(32) | 0.173 | 0.027-0.311 | 0.020 |
| LV/LA volume coupling indices: |  |  |  |  |  |
| LV<sub>ED</sub>/LA<sub>max</sub>, ratio | 1.79±0.69 | 1.55±0.56 | -0.191 | -0.328-(-)0.046 | 0.010 |
| LV<sub>dias</sub>/LA<sub>dias</sub>, ratio | 2.24±1.24 | 1.75±0.61 | -0.249 | -0.382-(-)0.107 | <0.001 |
| LV<sub>ED</sub>/LA<sub>min</sub>, ratio | 4.59±2.56 | 3.56±2.00 | -0.221 | -0.356-(-)0.076 | 0.003 |
| LV<sub>ES</sub>/ LA<sub>min</sub>, ratio | 1.75±1.04 | 1.36±0.77 | -0.209 | -0.346-(-)0.065 | 0.005 |
| LA function parameters: ‡ |  |  |  |  |  |
| Total Emptying Volume, ml | 33±17 | 36±19 | 0.076 | -0.071-0.219 | 0.308 |
| Total Emptying Fraction, % | 55±17 | 52±16 | -0.109 | -0.246-0.043 | 0.166 |
| Passive Emptying Volume, ml | 22±15 | 22±16 | -0.012 | -0.148-0.145 | 0.981 |
| Passive Emptying Fraction, % | 36±16 | 32±18 | -0.135 | -0.275-0.012 | 0.072 |
| Active Emptying Volume, ml | 11±9 | 14±11 | 0.142 | -0.004-0.282 | 0.057 |
| Active Emptying Fraction, % | 29±23 | 28±22 | -0.014 | -0.160-0.132 | 0.853 |
| Combined volume/Doppler indices: § |  |  |  |  |  |
| Indexed LA reservoir function<br>(LA total emptying fraction/$E/e'$), ratio | 0.082±0.054 | 0.056±0.023 | -0.312 | -0.457-(-)0.151 | <0.001 |
| Indexed LA conduit function<br>(LA passive emptying fraction/$E/e'$), ratio | 0.052±0.038 | 0.032±0.019 | -0.317 | -0.461-(-)0.157 | <0.001 |
| Diastolic LV dysfunction grade: \|\| |  |  |  |  |  |
| Normal | 54(65) | 58(59) | -0.196 | -0.350-(-)0.032 | 0.020 |
| Abnormal | 7(8) | 22(22) | 0.199 | 0.036-0.352 | 0.017 |
| Indeterminate | 22(27) | 17(17) | -0.109 | -0.251-0.038 | 0.147 |
| **B.** |  |  |  |  |  |
| LV mass, g | 123±35 | 130±38 | 0.088 | -0.064-0.236 | 0.255 |
| LV mass index, g/m<sup>2</sup> | 64±14 | 65±13 | 0.062 | -0.090-0.211 | 0.423 |
| LV<sub>dias</sub>, ml | 133±34 | 130±30 | -0.048 | -0.192-0.099 | 0.526 |
| LV<sub>dias</sub> index, ml/m<sup>2</sup> | 69±14 | 66±12 | -0.108 | -0.250-0.039 | 0.150 |
| LA<sub>dias</sub>, ml | 81±20 | 88±20 | 0.179 | 0.034-0.317 | 0.016 |
| LA<sub>dias</sub> index, ml/m<sup>2</sup> | 42±9 | 45±9 | 0.159 | 0.013-0.298 | 0.033 |
| LV<sub>dias</sub>/LA<sub>dias</sub>, ratio | 1.69±0.41 | 1.49±0.23 | -0.288 | -0.416-(-)0.148 | <0.001 |

\*, LV mass index female $\geq 95$ or male $\geq 115$ , g/m<sup>2</sup>; †, relative wall thickness ratio $>0.42$ and LV mass index female $<95$ or male $<115$ , g/m<sup>2</sup>; ‡, as per Blume et al.<sup>21</sup>; §, as per Ready et al.<sup>16</sup> with modification; ||, as per Naguen et al.<sup>20</sup> LV, left ventricular, LA, left atrial, LV<sub>ED</sub>, left ventricular end-diastolic volume, LV<sub>ES</sub>, left ventricular end-systolic volume, LV<sub>dias</sub>, left ventricular diastasis volume, LA<sub>max</sub>, left atrial maximum volume, LA<sub>dias</sub>, left atrial diastasis volume, LA<sub>min</sub>, left atrial minimum volume; E/e', early diastolic mitral inflow to mitral annular tissue velocities.

### CT coronary angiography

was acquired in a standard manner using 256iCT Philips scanner (Philips North America Corporation, Andover, MA). The iCT has 270-ms gantry rotation time and a temporal resolution of 135 ms. Scan parameters were 128X0.625 mm collimation. An intravenous injection of iodinated contrast media (Omnipaque 350) was used to opacify the coronary arteries and subsequently the LA and the LV. The scan was acquired during a single breath-hold. Scans were acquired with a Prospective ECG-Gated Axial Step-and-Shoot/Sequential technique. Only patients scanned in diastasis between 75%-81% of the R-R interval were chosen for the analysis with the standard acquisition occurring at 78%. LA volume was measured with exclusion for the LA appendage. LV volume was measured using an automated algorithm within the Cardiac software of the Philips Intellispace Portal. CT Diastolic Function Assessment – The diastolic expansion index was measured as ratio between diastasis LV and LA volume (LV_dias_/LA_dias_).^22^

### Statistical Analysis

Continuous variables are expressed as mean ± standard deviation and categorical variables are expressed as a number and percentages. Unpaired t-test, Chi-square, Fisher exact test or Meng’s Z-test for correlated correlation coefficients was used when appropriate for comparison between Group 1 and Group 2. For the predefined CT and TTE measurements, logistic regression analysis with the method of DeLong was used to assess the area under the receiver operating characteristic curve, Youden index J point with cut off value to differentiate patients from Group 2 and Group 1. Sensitivity and specificity were calculated in a standard manner. For all analyses, p values <0.05 were considered significant. All analysis were performed by JMP 15 (SAS Institute, Cary, NC, USA) and MedCalc Version 20.109 (Oostende, Belgium).

## RESULTS

Patient characteristics are presented in **Table 1**.

### Comparison of clinical data

Group 2 had higher blood pressure (mm Hg) for both systolic (138±15 *vs* 125±12, p<0.001, respectively), and diastolic (80±9 *vs* 75±9, p<0.001, respectively) readings. Hyperlipidaemia was noted more frequently in Group 2 (67 (69%) *vs* 44 (53%), p=0.027)

### Comparison of 2-Dimensional Echocardiographic Data (Table 2)

Patients in Group 2 had thicker interventricular septum (cm) (0.93±0.19 *vs* 0.85±0.16, p=0.005, respectively), LV posterior wall (cm) (0.87±0.17 *vs* 0.82±0.16, p=0.042, respectively), and mean LV wall thickness (0.90±0.16 *vs* 0.83±0.14, p=0.006, respectively). There was no presence of LV hypertrophy in either of the groups. The ratios of LV_ED_ to mean wall thickness as well as LV_ED_ index to mean wall thickness ratio were higher in Group 1 as compared to Group 2 (117±32 *vs* 107±29, p=0.028, respectively and 61±16 *vs* 55±14, p=0.005, respectively). The LV relative wall thickness ratio was higher in Group 2 (0.39±0.09 *vs* 0.35±0.08, p=0.004, respectively). Concentric LV remodelling was present more often in Group 2 (31 (32%) *vs* 12 (14%), p=0.020, respectively).

There were no differences between the groups in the measurements of the following: LV mass index, and LV ejection fraction. There were also no differences between the groups in the measurements of LV volumes (minimum - LV_ES_, maximum - LV_ED_, and in diastasis - LVdias).

The analysis of the LA showed that in Group 2, the LA was bigger for the following measurements: LA_max_ (67±26 *vs* 59±23, p=0.040, respectively), LA_dias_ (48±19 *vs* 41±17, p=0.006, respectively), and LA_dias_ index (25±9 *vs* 21±9, p=0.012, respectively), and both LA_min_ (31±14 *vs* 26±14, p=0.022, respectively) and LA_min_ index (16±7 *vs* 14±7, p=0.037, respectively).

All LV/LA volume coupling indices were lower in Group 2 as compared to Group 1, with the biggest difference being in diastasis: LV_dias_/LA_dias_ (1.75±0.61 vs. 2.24±1.24 p<0.001, respectively).

None of the standard LA function parameters differed between Group 1 and 2.

Indexed reservoir function (LA total emptying fraction divided by E/e’ ratio) and indexed conduit function (LA passive emptying fraction divided by E/e’ ratio) were higher in Group 1 (0.082±0.054 *vs* 0.056±0.023, p<0.001, respectively; and 0.052±0.038 *vs* 0.032±0.019, p<0.001).

### Comparison of Doppler Data (Table 2)

Group 2 had lower e’ values for both septum (6.8±1.7 cm/s *vs* 7.5±2.2 cm/s, p=0.047, respectively), and lateral wall (8.2±2.2 cm/s *vs* 9.7±3.2 cm/s, p=0.005, respectively). The mean value of E/e’ ratio averaged for both septum and lateral was higher in Group 2 (9.7±3.1 *vs* 8.1±2.8, p=0.003, respectively). There was no difference between Group 1 and Group 2 when the cut off value for E/e’ was set at above 14. Tricuspid Regurgitation velocity higher than 2.8 m/sec was more frequent in Group 2.

### Comparison of combined criteria for LV diastolic dysfunction (Table 2)

LV diastolic dysfunction was more common in Group 2. Normal LV diastolic function was noted in 58 (59%) patients in Group 2 and 54 (65%) patients in Group 1. LV diastolic dysfunction was noted in 22 (22%) patients in Group 2 and in 7 (8%) patients in Group 1. An indeterminate LV diastolic function was noted in 17 (17%) patients in Group 2 and in 22 (27%) patients in Group 1.

### CT Data (Table 2)

As noted in the TTE data, there was no difference between the groups for LV mass and LV mass index. There was no difference between the groups in LV volume measured in diastasis whether taken as mean value or indexed for body surface area. The ratio of LV_dias_/LA_dias_ was lower in Group 2 than in Group 1 (1.49 ±0.23 *vs* 1.68 ±0.41, p<0.001, respectively). Also, LA_dias_ was bigger in Group 2 for total volume (88 ±20 ml *vs* 81 ±20 ml, p=0.016, respectively), and indexed for body surface area (45 ±9 ml/m^2^ *vs* 42 ±9 ml/m^2^, p=0.033, respectively).

Receiver operating characteristics of both TTE and CT derived parameters are presented in **Table 3**. All measured parameters were analysed using receiver operating characteristics in order to discriminate patients with hypertension. We have followed the original concept of classifying the symptoms into one of the two categories, disease or no disease.

**Table 3.** Power analysis of (**A**) transthoracic echocardiographic and (**B**) computed tomography parameters for diagnosis of hypertension.

| Variable | Cut off<br>value | AUC | 95%<br>Confidence interval | Sensitivity<br>% | Specificity<br>% | Z statistics | P value |
| --- | --- | --- | --- | --- | --- | --- | --- |
| **A.** |  |  |  |  |  |  |  |
| Interventricular septal thickness, cm | >0.95 | 0.624 | 0.548-0.695 | 41 | 79 | 2.965 | 0.003 |
| Mean LV wall thickness, cm | >0.80 | 0.617 | 0.541-0.689 | 78 | 44 | 2.772 | 0.006 |
| LA<sub>max</sub>, ml | >52 | 0.592 | 0.517-0.665 | 69 | 48 | 2.169 | 0.030 |
| LA<sub>max</sub> index, ml/m<sup>2</sup> | >30 | 0.585 | 0.510-0.658 | 63 | 58 | 1.989 | 0.047 |
| LA<sub>dias</sub>, ml | >40 | 0.617 | 0.542-0.688 | 65 | 61 | 2.778 | 0.005 |
| LA<sub>dias</sub> index, ml/m<sup>2</sup> | >20 | 0.612 | 0.537-0.684 | 68 | 55 | 2.671 | 0.008 |
| LA<sub>min</sub>, ml | >21 | 0.622 | 0.547-0.693 | 78 | 46 | 2.896 | 0.004 |
| LA<sub>min</sub> index, ml/m<sup>2</sup> | >12 | 0.611 | 0.536-0.683 | 70 | 52 | 2.631 | 0.009 |
| e' lateral, cm/s | ≤9.5 | 0.643 | 0.546-0.732 | 75 | 57 | 2.633 | 0.009 |
| E/e' (average) | ≥8 | 0.662 | 0.576-0.741 | 70 | 59 | 3.394 | 0.001 |
| Tricuspid regurgitation, >2.8 m/sec | - | 0.556 | 0.467-0.643 | 16 | 95 | 2.141 | 0.032 |
| LV remodelling indices: |  |  |  |  |  |  |  |
| LV relative wall thickness, ratio | >0.40 | 0.626 | 0.550-0.697 | 40 | 82 | 3.013 | 0.003 |
| LV<sub>ED</sub>/mean wall thickness | ≤124 | 0.604 | 0.528-0.677 | 75 | 43 | 2.432 | 0.015 |
| LV<sub>ED</sub> index/mean wall thickness | ≤57 | 0.627 | 0.551-0.698 | 61 | 62 | 3.007 | 0.003 |
| Concentric remodelling |  | 0.572 | 0.496-0.645 | 29 | 86 | 2.386 | 0.017 |
| LV/LA volumetric coupling indices: |  |  |  |  |  |  |  |
| LV<sub>dias</sub>/LA<sub>dias</sub> | ≤1.81 | 0.627 | 0.552-0.698 | 65 | 58 | 3.020 | 0.002 |
| LV<sub>ES</sub>/LA<sub>min</sub> | ≤1.23 | 0.614 | 0.538-0.686 | 54 | 67 | 2.686 | 0.007 |
| LV<sub>ED</sub>/LA<sub>min</sub> | ≤3.81 | 0.629 | 0.554-0.700 | 70 | 55 | 3.051 | 0.002 |
| LV<sub>ED</sub>/LA<sub>max</sub> | ≤1.24 | 0.626 | 0.550-0.697 | 38 | 87 | 3.008 | 0.003 |
| Combined volumetric/Doppler indices: * |  |  |  |  |  |  |  |
| Indexed LA reservoir function<br>(LA total emptying fraction/<sub>E/e'</sub>), ratio | ≤0.068 | 0.694 | 0.609-0.770 | 74 | 62 | 4.103 | <0.001 |
| Indexed LA conduit function<br>(LA passive emptying fraction/<sub>E/e'</sub>), ratio | ≤0.053 | 0.678 | 0.592-0.756 | 92 | 44 | 3.726 | <0.001 |
| Diastolic LV dysfunction grade: † |  |  |  |  |  |  |  |
| Normal | - | 0.580 | 0.494-0.663 | 27 | 89 | 2.468 | 0.014 |
| Abnormal | - | 0.581 | 0.495-0.663 | 27 | 89 | 2.512 | 0.012 |
| **B.** |  |  |  |  |  |  |  |
| LA<sub>dias</sub>, ml | >77 | 0.617 | 0.542-0.689 | 66 | 59 | 2.766 | 0.006 |
| LA<sub>dias</sub> index, ml/m<sup>2</sup> | >44 | 0.602 | 0.527-0.674 | 52 | 67 | 2.411 | 0.016 |
| LV<sub>dias</sub>/LA<sub>dias</sub>, ratio | ≤1.76 | 0.643 | 0.569-0.713 | 91 | 36 | 3.381 | <0.001 |

\*, as per Ready et al.<sup>16</sup> with modification; †, as per Naguen et al.<sup>20</sup>; LV, left ventricular; LA, left atrial; LV<sub>ED</sub>, left ventricular end-diastolic volume; LV<sub>ES</sub>, left ventricular end-systolic volume; LV<sub>dias</sub>, left ventricular diastasis volume; LA<sub>max</sub>, left atrial maximum volume; LA<sub>dias</sub>, left atrial diastasis volume; LA<sub>min</sub>, left atrial minimum volume; E/e', early diastolic mitral inflow to mitral annular tissue velocities; AUC, area under the curve.

Predictive modelling for **CT data** showed that the following measurements had discriminatory power to differentiate patients with hypertension: (i) for LA_dias_ >77 ml (area under the curve (AUC) 0.617, confidence interval (CI) 0.542-0.689, p=0.006); (ii) for LA_dias_ index >44 ml/m^2^ (AUC 0.602, CI 0.527-0.674, p=0.016), (iii) and for LV_dias_/LA_dias_ ratio ≤1.76 (AUC 0.643, CI 0.569-0.713, p<0.001). The LV_dias_/LA_dias_ ratio ≤1.76 had the highest sensitivity of 91% but low 36% specificity.

Predictive modelling for **TTE data** includes a few parameters listed in **Table 3**. The three parameters with AUC >0.63 were: E/e’ (average) ≥8 (AUC 0.662, CI 0.576-0.741, p=0.001), e’ lateral ≤9.5 (AUC 0.643, CI 0.546-0.732, p=0.009), and LV_ED_/LA_min_ ≤3.81 (AUC 0.629, CI 0.554-0.700, p=0.002) (**Figure 2A**). All three had similar range of sensitivity and specificity. The two indexed LA function indices were lower in hypertension (**Figure 2B**): indexed reservoir function (AUC 0.694, CI 0.609-0.770, p<0.001), and indexed LA conduit function (AUC 0.678, CI 0.592-0.756, p<0.001). Their sensitivity and specificity were 74%, 62% and 92% and 44%, respectively.

**Figure 2.**
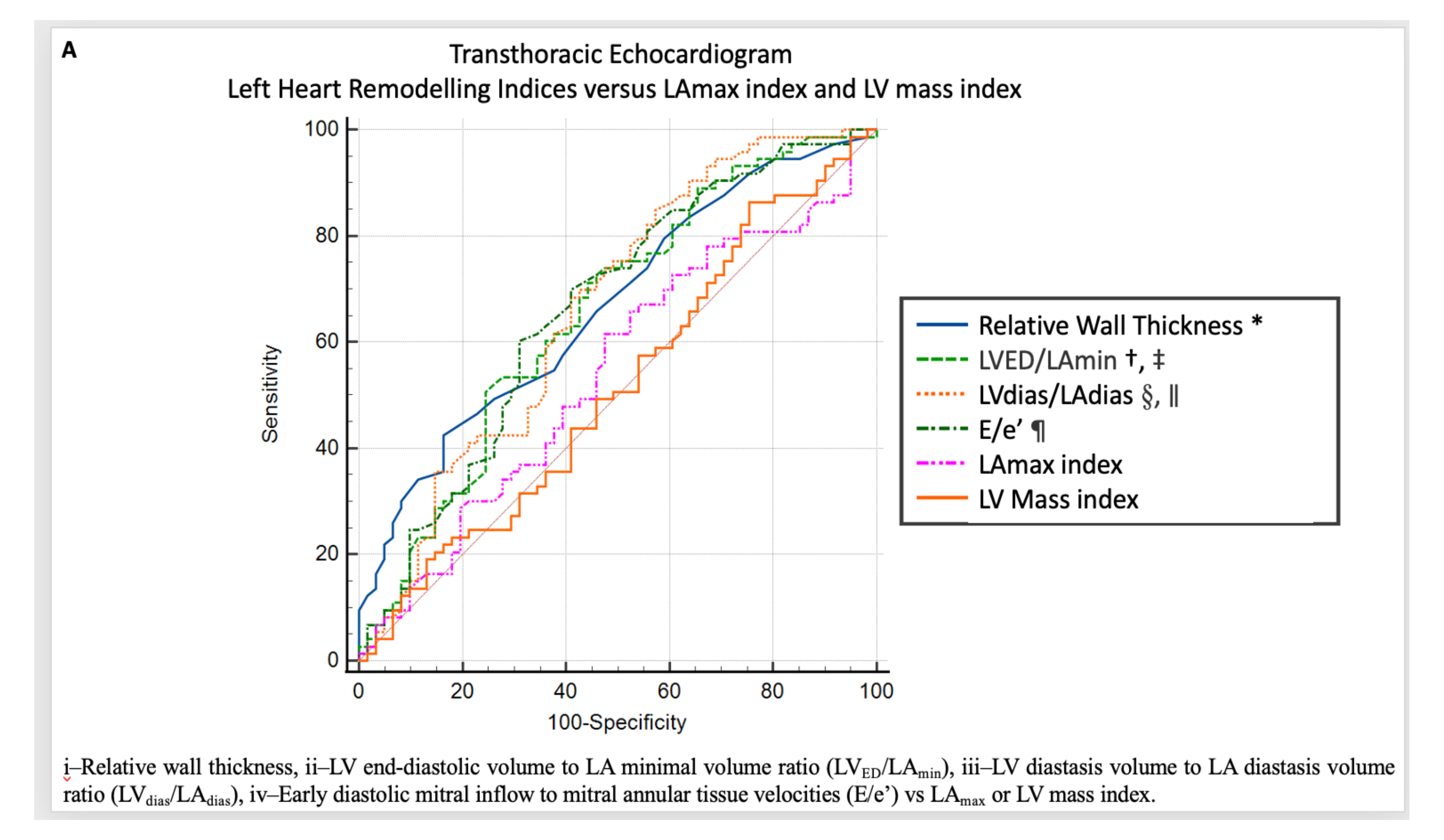
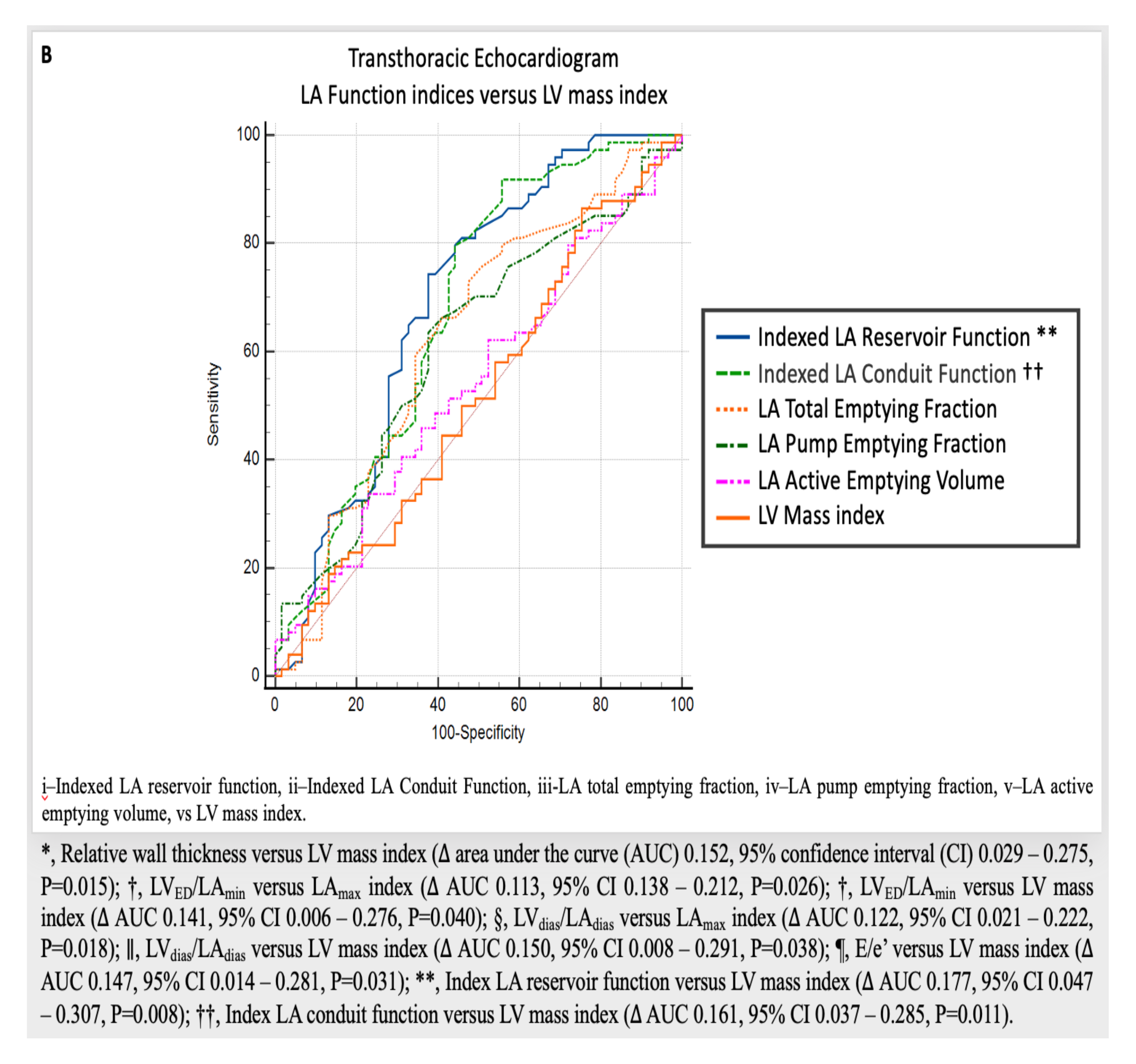
Comparison between the receiver operating characteristics curves for echocardiographic parameters for diagnosis of hypertension, **A**-left heart remodelling indices; **B**-Left atrial (LA) function as compared to left ventricular (LV) mass index and/or left atrium maximum volume (LA_max_) index.

### Parameters that have the best predictive value for the presence of hypertension (Figure 3)

Three TTE derived parameters were chosen for their best overall accuracy to predict the presence of hypertension. These are: 1- relative wall thickness >40, 2– LV_dias_/LA_dias_ ≤1.81, and 3–LA reservoir function indexed by E/e’≤0.068. They were subsequently grouped together to propose an overall diagnostic score. This method provided the highest AUC of 0.737. Sensitivity, specificity, and overall accuracy of combined TTE score are shown in **Table 4**. The score was graded accordingly where 0 is the minimum, and 3 is the maximum score obtained if each of the three parameters scored one point.

**Figure 3.**
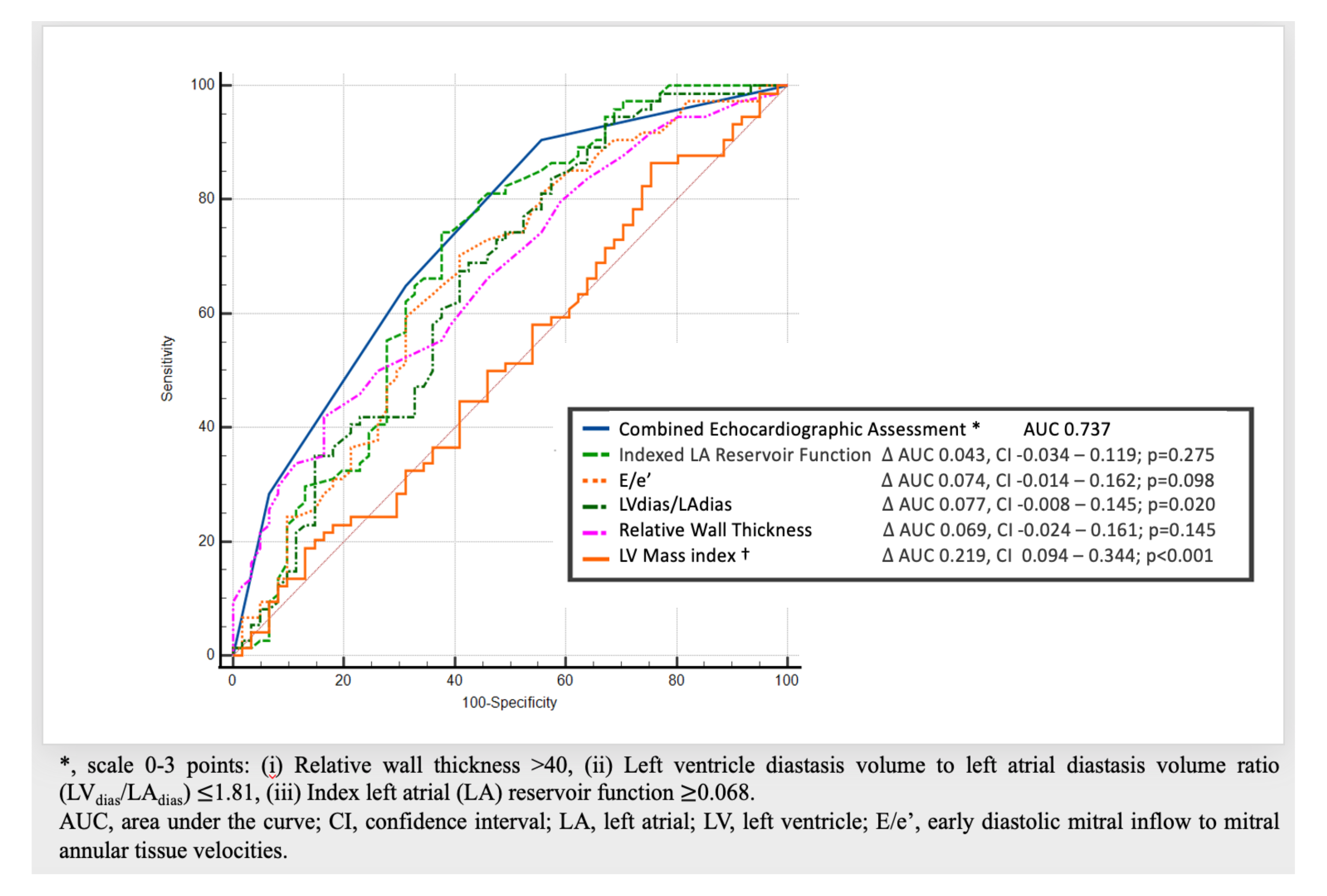
Power analysis of echocardiographic parameters to diagnose hypertension.

**Table 4.** Sensitivity and specificity of combined echocardiographic parameters scoring to diagnose patients with hypertension.

| Score* | Sensitivity | 95% | Specificity | 95% |
| --- | --- | --- | --- | --- |
|  | % | confidence interval | % | confidence interval |
| 0 | 100 | 96-100 | 0 | 0-4.3 |
| 1 | 84 | 75-90 | 45 | 34-56 |
| 2 | 56 | 45-66 | 75 | 64-84 |
| 3 | 22 | 14-31 | 95 | 88-99 |

\*, Score from 0-3. One point for each: (i) relative wall thickness $>40$ , (ii) left ventricular diastasis volume to left atrial diastasis volume ratio $(LV_{dias}/LA_{dias}) \leq 1.81$ , and (iii) indexed by $E/e'$ (early diastolic mitral inflow to mitral annular tissue velocities) LA reservoir function $\leq 0.068$ .

## DISCUSSION

The presence of LV hypertrophy remains the hallmark of hypertension and represents the effect of the maladaptive process that leads to contractile dysfunction.^6, 11, 12, 23^ It is the most widely used measure of assessment for established features of systemic hypertension and is familiar to clinicians. The sequence of gradual adaptive changes in left heart anatomy and function that eventually lead to the presence of LV hypertrophy is of ongoing interest.^7, 8, 24, 25^ Most of the previous studies which describe noted features of early hypertension are predominantly based on studying the changes isolated either to the LV and/or to the LA. In this study the concept of volume ratio as a single left atrioventricular parameter in detecting early remodelling in hypertension was assessed. By using TTE we were able to compare the diagnostic values of preselected phases of the cardiac cycle. The concept of the ratio of LA to LV diameter as a potential non-invasive marker of LV compliance was used previously using TTE, and LA to LV diameter ratio ≥1.0 was observed in hypertension and diabetes.^15^ The importance of left atrioventricular coupling index was highlighted by Pezel et. al.^26^, who derived the ratio of LA to LV volume, from end-diastole, using Cardiac Magnetic Resonance. They have noted that the ratio is a strong predictor for the incidence of HF, atrial fibrillation, and coronary disease mortality.^26^ Since the cumulative effect of hypertension leads to cardiac remodelling and vascular aging^27^, early diagnosis is paramount. Therefore, establishing a protocol that is easy to follow, of routinely obtained parameters, to detect and to monitor hypertension, would be very useful in clinical routine in improving the diagnosis rate and to improve long term prognosis.

In this patient population of hypertensive patients, we found that there was a trend among left heart coupling indices, all of them being available from a routine clinical echocardiogram, each of which allowed to bring new information on left heart remodelling process in hypertension.

### LA Volume and Function

Larger LA volumes were noted at end-systole - maximum (LA_max_), end-diastole - the minimum (LA_min_), and in diastasis (LA_dias_). This process of LA dilatation is a response to increasing LV filling pressure, in due course increasing LA pressure and finally, increased LA wall stress.^25, 28^ In order to maintain adequate preload in the setting of increasing LV filling pressure, LA contraction will need to increase correspondingly.^29^ We have observed only a borderline increase in active emptying volume (p=0.057), and a borderline reduction in passive LA emptying. However, when total emptying fraction (reservoir) and passive emptying fraction (conduit) were indexed by E/e’ ratio, we found those to be lower in patients with hypertension. This finding is in support of the concept that the changes in LA reservoir and LA conduit dysfunction are noted earlier and LA systolic dysfunction occurs later in disease progression and is secondary to LV structural remodelling and LA afterload mismatch.^24, 30^ LA diastolic filling i.e. reservoir filling, is determined by both LA relaxation and compliance, the latter being determinant of stroke volume and an important determinant of LA function.^31^ A similar reduction in indexed LA reservoir function was noted in the past in patients with HF with preserved ejection fraction.^16^ In the current study, indexed LA reservoir function and indexed LA conduit function were found both to be better than LV mass index in diagnosing hypertension with their cut off values of 0.068 and 0.053, respectively.

### Volume ratio Analysis

Earlier studies of others used retrospective cardiac CT volumetric data in the evaluation of diastolic dysfunction and prediction of HF.^32^ In this study, prospective gated cardiac CT diastasis volumetric data and TTE were used to analyse LV/LA volume ratio in selected phases of the cardiac cycle. Despite the absence of both LV dilatation and LV hypertrophy, the ratio of LV/LA volume was reduced in patients with hypertension. This LV/LA volume ratio index was reduced not only when measured in diastasis (LV_dias_/LA_dias_) using both CT and TTE, but also in TTE as maximum to minimum (LV_ED_/LA_min_), maximum to maximum (LV_ED_/LA_max_), and minimum to minimum (LV_ES_/LA_min_). Power analysis of TTE derived volume ratio indices showed that LV_ED_/LA_min_ ratio and LV_dias_/LA_dias_ were the most accurate to distinguish patients with hypertension with cut off values of 3.81 and 1.81, respectively. Both were more accurate in describing hypertension than either LA_max_ index or LV mass index.

The three TTE parameters with AUC>0.63 were: E/e’ (average) ≥8, e’ lateral ≤9.5, and LV_ED_/LA_min_ ≤3.81. All three had a similar range of sensitivity and specificity. The advantage of LV/LA volume ratio is that it is a simple parameter, available in most patients and as examined in this paper, of value using both TTE and CT coronary angiography. Whether this will be also applicable to other cardiac imaging techniques remains to be seen.

### Combined TTE Scoring Index

We have proposed a new scoring index in which three parameters: (1) relative wall thickness > 40; (2) LV_dias_/LA_dias_ ≤ 1.81; and (3) indexed LA reservoir function ≤0.068, are used to diagnose patients with hypertension. Each parameter if present is worth one point and the maximum total score of three is possible. This concept of combining those three parameters proved more accurate than using either LV mass index, relative wall thickness, or any of the other measured parameters alone. The score of 3 was 95% specific with the score of 2 showing 75% specificity and 56% sensitivity to diagnose hypertension. The score of 0 to 1 had high sensitivity of 100% to 84% but low specificity to exclude hypertension.

## PERSPECTIVES

Using TTE and CT data, a new left atrioventricular coupling index was identified to detect early left heart remodelling in patients with hypertension. An abnormal ratio of LV to LA volume measured in diastasis, using both TTE and CT, was a predictor of underlying hypertension. A combined power of three TTE parameters had a better diagnostic value than individual LA or LV parameters measured separately. The new index score of three parameters could prove useful in improving early diagnosis, define progress of left heart remodelling, and subsequently may lead to the improvement of long term prognosis.

## Data Availability

on request

## Abbreviations and non-standard acronyms

CT: Computed Tomography
HF: Heart Failure
LA: Left Atrial/Atrium
LA_dias_: Left Atrial diastasis volume
LA_max_: Left atrial maximum volume
LA_min_: Left atrial minimal volume
LV: Left Ventricle/Ventricular
LV_dias_: Left Ventricular diastasis volume
LV_ED_: Left Ventricular end-diastolic volume
LV_ES_: Left Ventricular end-systolic volume
TTE: Transthoracic Echocardiogram

## Acknowledgments

We would like to thank Professor Robert Ware from the Griffith University, Queensland for his critical comments and statistical advice. We would like to thank Professor Andrzej Lange for his critical comments and guidance.

## Sources of Funding

This project was supported by Wesley Medical Research Clinical Research Grant #2020-23.

## Disclosures

All authors report that they have no relationship with the industry to be disclosed.

## Notes

### Competing Interest Statement

The authors have declared no competing interest.

### Author Declarations

The study was approved by UnitingCare Health Human Research Ethics Committee (number 2019.29.307), Brisbane, Australia.

